# WT1 Tumor Antigen is Overexpressed in Kaposi Sarcoma and is Regulated by KSHV vFLIP

**DOI:** 10.1101/2021.02.20.21251636

**Authors:** Ayana Morales, Caitlyn Genovese, Matthew Bott, Julio Alvarez, Sung Soo Mun, Jennifer Totonchy, Archana Gautam, Jesus Delgado de la Mora, Stephanie Chang, Maite Ibáñez de Garayo, Dagmar Wirth, Marcelo Horenstein, Tao Dao, David A. Scheinberg, Paul G. Rubinstein, Aggrey Semeere, Jeffrey Martin, Margaret Borok, Thomas B. Campbell, Susan E. Krown, Ethel Cesarman

## Abstract

**Purpose:** Wilms’ tumor 1 (WT1) is overexpressed in several cancers, and WT1 expression levels are associated with poor prognosis. As a host protein that functions as an oncogene, it represents an important immunotherapeutic target. This study evaluated WT1 expression in Kaposi sarcoma (KS) tumors to assess whether immunotherapy targeting WT1 is a potential therapeutic approach for KS. We also investigated the role of the causal agent of KS, Kaposi sarcoma herpesvirus (KSHV/HHV-8) in regulating WT1 expression.

**Experimental design:** Immunohistochemistry for WT1, KSHV, and B and T cells subsets, followed by image analysis, was performed in 363 KS tumor biopsies. Expression of KSHV vFLIP was evaluated by immunofluorescence. Primary endothelial cell cultures and cell lines were infected with KSHV *in vitro*, or transduced with an inducible vFLIP vector and induced with doxycycline, and then assessed for WT1 expression. Binding of ESK-1, a T cell receptor mimic therapeutic antibody that recognizes WT1 peptides presented on MHC HLA-A0201, was assessed using flow cytometry.

**Results:** We report overexpression of WT1 in KS tumors, which was associated with increased with increasing histopathologic stage and the proportion of KSHV-infected cells. Areas with high WT1 expression showed sparse T cell infiltrates. KSHV infection *in vitro* resulted in WT1 upregulation, mediated by the viral protein vFLIP, which resulted in stronger binding of ESK1.

**Conclusions:** KS lesions express high levels of WT1, a process regulated by the KSHV-encoded vFLIP. These findings suggest that immunotherapy directed against WT1 may represent a therapeutic approach for this cancer.

**Translational Relevance:** Kaposi sarcoma (KS) is a vascular neoplasm caused by the Kaposi sarcoma herpesvirus (KSHV/HHV-8). People living with HIV are not only at a significantly higher risk of developing KS, but also often have a more aggressive clinical course. Although antiretroviral therapy may cause regression of HIV-associated KS lesions, advanced cases of KS also require chemotherapy, which is rarely curative. Wilms’ tumor 1 (WT1) has been reported to be overexpressed in various cancers, functioning as an oncogene and associated with a poor prognosis. WT1 is also an important immunotherapeutic target, with several WT1-directed therapies showing promising results in early clinical trials for leukemias and solid tumors. Here we report high expression of WT1 in KS, especially in higher histological stages. Our findings provide pre-clinical evidence that supports conducting anti-WT1 immunotherapy trials in KS, and evaluating WT1 expression as a potential biomarker to identify individuals most likely to benefit.

## Introduction

Kaposi sarcoma herpesvirus (KHSV), also called human herpesvirus 8 (HHV-8), the etiologic agent of Kaposi sarcoma (KS), has been classified as a carcinogen by the International Agency for Research on Cancer (IARC).^1^ KSHV viral DNA and protein are detected in all KS tumors^2, 3^. KS is a vascular neoplasm that may present as a few lesions confined to the skin, but may progress to multiple lesions that can involve the oral mucosa, lymphatics, lungs and other visceral organs^3^. Morphologically, KS lesions may present as patches, plaques and nodules. Despite effective antiretroviral therapy, KS remains the most common cancer in people living with human immunodeficiency virus (HIV) globally. While KS also occurs in people without HIV infection, HIV/AIDS associated KS is generally the most aggressive form, occurring most often in HIV- and KSHV-co-infected individuals in sub-Saharan Africa and elsewhere in men who have sex with men (MSM)^4-6^. People living with HIV (PLWH) may have a up to a 500-fold excess relative risk for the development of KS compared to the general population^5-7^.

Furthermore KS continues to occur in PLWH despite HIV infection that is well controlled with antiretroviral therapy (ART)^8^. While ART alone may lead to tumor regression in limited-stage KS, chemotherapy is often required for patients with advanced KS, ^4, 9-11^ is rarely curative, and incurs a risk for severe toxicities^12^. While immunosuppression plays a role in KS progression, there are additional viral and host factors involved. The contribution of virus and host factors to the molecular pathogenesis of KS may identify potential targets of therapy and diagnostic as well as prognostic biomarkers.

Wilms’ Tumor 1 (WT1) is a C2H2 zinc-finger transcription factor that was initially discovered as a tumor suppressor. However, when overexpressed, it has been recognized as promoting carcinogenesis in various hematological malignancies and solid tumors^13, 14^. Wild type WT1 is expressed in isolated sites in adult tissues, (e.g. the glomerular podocytes of the kidney, the sertoli/granuloma cells of the testis/ovary and 1% of bone marrow cells., and the mesothelium surrounding visceral organs^15^), but is frequently overexpressed in human leukemias and solid tumors, including lung, brain, esophageal and ovarian cancers, as well as Wilms’ tumors, and exhibits pro-oncogenic functions that are associated with a worse prognosis^13, 14, 16^. Differing functions of WT1 are a result of at least 36 different isoforms resulting from alternative splice variants and from different translation initiation sites, including the pro-tumorigenic (cug) or tumor suppressive (aug) isoforms, as well as posttranslational modifications^17^. The cugWT1 N-terminally phosphorylated isoform is overexpressed in many cancers, induces cellular transformation, and increases target gene expression of C-MYC, BCL-2, and EGFR^17^. Wild-type WT1 overexpression is associated with increased cell proliferation, survival, angiogenesis, epithelial to mesenchymal transformation, and inhibition of apoptosis^18-20^.

In 2009, the National Cancer Institute (NCI) ranked WT1 as the number one cancer antigen toward which to direct immunotherapy research based on (1) therapeutic function, (2) immunogenicity, (3) oncogenicity, (4) specificity, (5) expression level, (6) stem cell expression, (7) number of antigen positive cancers, (8) antigenic epitopes, and (9) cellular location of antigen expression^21^. Immunotherapy targeted against WT1 in various hematological malignancies and solid cancers has induced immunological and clinical responses in preclinical studies^22^ and clinical trials^23-25^. In particular, ESK-1, a T cell receptor mimic antibody targeted against the RMF peptide of WT1 and the HLA-A0201 antigen, has shown activity against leukemia and solid tumor cell lines in preclinical animal models ^22^. WT1-directed immunotherapy in the form of peptide vaccines has been studied in Phase 1 and 2 clinical trials in acute myelocytic leukemia (AML)^25^, malignant pleural mesothelioma^26^, and multiple myeloma (ClinicalTrials.gov, NCT01827137) and has shown safety and clinical and immunological responses. WT1 vaccination is being studied in combination with immune checkpoint inhibitors (anti-PD-1) in patients with multiple relapsed or refractory solid tumors and leukemias (ClinicalTrials.gov, NCT03761914). Recently, donor derived, EBV specific CD8+T cells with high affinity WT1 specific TCR (TCR_C4_) HLA-A2 matched, generated persistent T cell responses that prevented relapse in 100% of 12 patients with high risk AML compared to 88 patients with similar risk AML with 54% relapse free survival^27^. In addition, WT1 serves as a prognostic marker in AML, myelodysplastic syndromes and solid tumors^13, 14^.

Like other herpesviruses, KSHV can establish lifelong latency, and in KSHV-induced tumors, the majority of cells are latently infected^3^. KSHV expresses a handful of latent viral oncoproteins, including LANA, vCyclin and vFLIP, which are thought to drive cellular transformation^3, 28^. In particular, the latent gene vFLIP, induces vascular proliferation, spindle cell morphology and an inflammatory phenotype when expressed in endothelial cells. vFLIP activates IκB kinase 1 (IKK1) to stimulate nuclear factor κB (NFκB) signaling to increase cell survival by directly binding to IKK*γ* (NEMO)^29-32^. There are putative NFκB binding sites within the WT1 promoter, and ectopic expression of p50 and p65 subunits of NFκB upregulate WT1 promoter activity^33^. One study reported analysis of vFLIP ectopic expression on human umbilical vein endothelial cells (HUVEC) followed by cDNA microarray analysis, and WT1 RNA was among a number of upregulated transcripts^34^.

WT1 expression was examined in a variety of vascular tumors, including a few cases of non-HIV-associated KS, and expression was reported in 8/19 cases^35, 36^. However, the spectrum of WT1 expression in KS is not well studied, and neither the mechanism of WT1 upregulation nor the spatial relationship of WT1 and immune infiltrates in KS have been previously investigated. Hence, we examined WT1 expression in a large cohort of HIV-associated KS, plus additional cohorts of both non-HIV- and HIV-associated KS. We also investigated whether KSHV plays a direct role in WT1 expression.

## Materials and Methods

### Patient specimens

KS biopsies were obtained from several cohorts. The largest comprised of 334 KS tumor biopsies obtained as part of a joint AIDS Malignancy Consortium (AMC) and AIDS Clinical Trials Group (ACTG) clinical trial, AMC066/A5263 (NCT01435018), collected at 11 sites in sub-Saharan Africa and Brazil, with approval from local and national ethics committees^37^. A total of 303 specimens of KS biopsies were evaluable. This clinical trial included adults living with HIV who had advanced, biopsy confirmed KS, and who had not previously received local or systemic chemotherapy or radiotherapy^37^. Participants were enrolled between 2013 and 2018 in an open label, non-inferiority trial examining optimal treatment regimens for AIDS associated KS in resource limited areas with high KS prevalence^37^. A second cohort comprised 20 KS biopsies from PLWH from the Infectious Disease Institute of Makerere University in Uganda. A third cohort comprised 10 KS biopsies from patients with HIV-associated KS from Stroger Cook County Hospital in Chicago. A fourth cohort included 30 KS biopsies from the Department of Pathology and Laboratory Medicine of Weill Cornell/New York Presbyterian Hospital, half of which were PLWH, and the remainder had no clinical evidence of HIV infection or iatrogenic immunosuppression. Approval was obtained from the respective institutional IRBs for use of all tissue specimens.

### Cells and reagents

iSLK cells that harbor wild-type KSHV BAC16^38, 39^ were used to make KSHV virus stocks (gift from Ashlee Moses) and were cultured in Dulbecco’s modified Eagle’s medium (DMEM) (GIBCO) with 10% FBS, 1% gentamicin sulfate (Caisson Labs), 1 µg/ml puromycin (Thermo Fisher Scientific), 250 µg/ml G418 (Thermo Fisher Scientific) and 1.2 mg/ml hygromycin B (Corning). HuARLT-1 cells, a conditionally immortalized human endothelial cell line^40^ carrying doxycycline-dependent cassettes for autoregulated expression of the SV40 Tag and hTert, were cultured in endothelial cell growth medium EGM-2 (Lonza) with 2µg/ml of doxycycline. Human umbilical vein endothelial cells (HUVEC) (Lonza) were cultured in EGM-2. iLEC cells were generated as previously described^41^ and were cultured in EMG-2-MV (Lonza). TIVE (telomerase-immortalized human umbilical vein endothelial) and TIVE-LTC (long term-infected TIVE) cells (gift from Rolf Renne, Department of Molecular Genetics and Microbiology, University of Florida) were cultured in EGM-2. Human embryonic kidney 293T cells (HEK293T cells) (ATCC) were cultured in DMEM with 10% FBS, 1% gentamicin sulfate. Doxycycline hyclate (Sigma-Aldrich) was used at 1-2µg/ml for induction of vFLIP.

### Plasmids and antibodies

WT vFLIP and vFLIP NFκB dead mutant inducible cell lines were established using previously described doxycycline inducible lentiviral vectors^42^. Stable transduced cell lines were established in HUVEC and HuARLT-1 by puromycin selection at 1µg/ml and doxycycline 1-2µg/ml for inducible expression of WT vFLIP and vFLIP null NFκB. HuARLT-1 cells were then transduced with a gamma retroviral vector with HLA-A0201 packaged from 293T HEK. Stable HuARLT-1 cell lines expressing HLA-A0201 were sorted using an ARIA-2 flow cytometer within the Weill Cornell Medicine Cytometry Core.

### KSHV BAC-16 Infection

KSHV wild type BAC-16 stocks of virus were prepared from stable iSLK cells as previously described^39^. iSLK-BAC-16 were cultured in DMEM supplemented with 10% FBS, 1% gentamicin sulfate, 1 µg/ml puromycin, 250µg/ml G418 and 1,2000 µg/ml hygromycin B. Stable iSLK-BAC16 cells were induced with the addition of both doxycycline (1 μg/ml) and sodium butyrate (1 mM). Four days later, supernatant was collected and cleared of cells and debris by centrifugation (2,000 rpm for 5 min at 4°C) and filtration (0.45 μm). Virus particles were pelleted by ultracentrifugation (25,000 rpm for 1.5 h at 4°C) using an SW32 Ti rotor. A spinoculation with KSHV was performed on the respective cell line, using KSHV virus in the presence of 8µg/ml polybrene (Sigma-Aldrich) in serum-free media, at 1000rpm for 30 min at 4°C. Images were obtained at 20x using the Olympus BX63 Fluorescence Microscope with DP80 Camera.

### Lentiviral Transduction

Lentiviral packaging was performed using Effectene (QIAGEN) per packing instructions with the packaging plasmids psPAX2 and vsvg into HEK293T cells. The WT1 shRNA and control plasmids were obtained from (Dharmacon). Supernatant containing lentivirus particles were collected at 48 and 72 hours and filtered (0.22 μm) and stored at −80°C. HUVEC, HuARLT-1 cells, and 293T cells underwent spinoculation, with filtered supernatant in the presence of 8µg/ml polybrene (Millipore-Sigma) in serum free media, at 1000rpm for 30 min at 4°C. Transduced cells were then selected using puromycin 1µg/ml.

### Interferon gamma treatment

HuARLT-1 cells were treated with IFN-*γ* (recombinant human IFN-*γ*) (R&D Systems) at 50ng/ml for 72 hours. Cells were then harvested for protein for western blotting.

### Western blotting

Total protein extracts were prepared from *in-vitro* 2D cell culture systems using RIPA buffer (Thermo-Fisher). Quantification of protein lysates was done using Pierce BCA assay (Pierce). Proteins were separated using 10% sodium dodecyl sulfate-polyacrylamide gel electrophoresis SDS-PAGE gel (BioRad) and then transferred to a polyvinylidene difluoride membrane (PVDF) (GE Healthcare) and blocked in 5% nonfat dry milk-TBST for 1 hour at room temperature. The PVDF membrane was then incubated overnight with primary antibodies diluted in 5% BSA-TBST overnight at 4°C and then immunoassayed using standard methods. The following antibodies were used for western blotting: mouse monoclonal antibody WT1 clone F-6, (1:200, Santa Cruz Biotechnology), mouse monoclonal WT1-NT (clone 6F-H2, EMD Millipore), Anti-Flag (1:1000, Rockland). A rat monoclonal antibody to vFLIP (1:200, clone 4C1) was kindly provided by Elisabeth Kremmer at Helmholtz Zentrum Munchen, Germany. Secondary anti-HRP rabbit antibody (1:5000, GE healthcare), secondary anti-HRP mouse antibody (1:2000, GE healthcare) and goat anti-rat IgG (H&L) HRP antibody (1:5000, Thermo Fisher Scientific) were used for western blotting.

### qPCR

Total RNA was isolated according to Qiagen RNAeasy Mini kit (QIAGEN) standard protocols. RNA purity was confirmed with absorbance at 260nm. The reverse transcription reaction was adapted from protocols supplied by Applied Biosystems. Briefly, reactions were incubated for 10 minutes at 95°C followed by 50 cycles of 15 seconds at 95°C and 60 seconds at 60°C. All reactions were completed in triplicate. The quantitative RT-PCR and fluorescence measurements were made using the Applied Biosystems 7500 Real-Time PCR System. B-actin and GAPDH were utilized as housekeeping genes for normalization. Primers used for qPCR are listed in **supplemental Table 1**.

### Histopathological classification and immunohistochemistry (IHC)

All cases were reviewed by two pathologists (MH and EC) to confirm the presence of KS and were classified into one of three histologic stages (patch, plaque and nodule) as described^3^. Immunophenotyping was performed on formalin-fixed, paraffin-embedded tissue sections on a Leica Bond III system using the standard protocol. Sections were pre-treated using heat-mediated antigen retrieval with Sodium-Citrate buffer (pH6, epitope retrieval solution 1) for 30 mins. The sections were then incubated with appropriate antibodies for 15 mins at room temperature and detected using an HRP-conjugated compact polymer system. 3,3’-Diaminobenzidine (DAB) was used as the chromogen. Sections were then counterstained with hematoxylin and mounted with micromount. Mouse monoclonal 6F-H2 WT1 (DAKO) and anti-LANA rat monoclonal HHV-8 ORF72 clone LN53 (Abcam) were used for IHC. T cell subsets were assessed by IHC with mouse monoclonal CD4 (clone 4B12, Leica) and mouse monoclonal CD8 (clone 4B11, Leica).

### HALO Analysis to quantify Immunohistochemistry staining

Immunostained slides for WT1 and LANA were scanned at 20x (Aperio, Leica Biosystems). Using HALO Imaging analysis software, WT1 and LANA staining was quantified as percent positive cells in areas involved by KS or adjacent uninvolved skin. WT1 was graded by intensity based on percentage of positively stained cells, 1-20% (1+), 21-50% (2+), >50% (3+). Correlation analysis was assessed for WT1 and LANA. HALO analysis was also performed for CD4+T cells and CD8+T cells, staining by percentage within areas involved by KS and surrounding stromal tissue. Correlation of WT1 and T cell subsets (CD8 and CD4) cells was assessed in all evaluable cases. A subset of cases with available sequential sections were examined to determine whether there was a spatial correlation of LANA, WT1 and immune infiltrates (CD4+ and CD8+ T cells). These included 13 nodules, 11 plaques and 12 patch lesions. Five independent areas with high LANA or high CD8 cell numbers, were analyzed in the corresponding adjacent section for other markers (LANA, CD8, CD4 and WT1). The number of cells and percentage of LANA+, CD4+ and CD8+ cells were quantified using HALO software with analysis tools, “Image Registration” and “Synchronization”. The total number of WT1+ cells, CD4+T cells, and CD8+T cells were quantified in LANA-rich areas. Conversely, five high CD8+ regions were selected and the total number of LANA+, CD8+, CD4+, and WT1+ cells were quantified in sequential sections. Proportional values of all indices were evaluated in both high LANA+ sections and high CD8+ T cells on corresponding sections. These percentages were calculated and plotted using Graph Pad Prism version 8.0 for Windows (GraphPad Software, San Diego, CA, USA).

### Immunofluorescence on primary KS tissue

KS tissue sections were deparaffinized by melting at 60°C for 10 minutes followed by 10 minute incubation in xylene. Slides were rinsed once with xylene, twice with 100% ethanol, and three times with water. Antigen retrieval was performed with sodium-citrate buffer (pH6, epitope retrieval solution 1) for 10 minutes at 37°C in a humidity chamber. Slides were rinsed three times with water and peroxidase inhibition was performed with 30% H_2_O_2_ for 10 minutes at room temperature. After three rinses with PBS, slides were blocked in 5% normal goat serum in PBS for 15 minutes in a humidity chamber. Slides were rinsed again three times with PBS before and between the following steps: incubation with primary antibody rat anti-vFLIP-4C1 at 1:5 dilution in 2% TSA block (Invitrogen) for three hours at room temperature in a humidity chamber; incubation with anti-rat HRP secondary at 1:100 dilution and DAPI at 1:1000 dilution in 2% TSA block for 30 minutes at room temperature in a humidity chamber; incubation with AlexaFluor-594 tyramide at 1:100 in amplification buffer containing 0.05% H2O2 for 10 minutes in a humidity chamber; and mounting of coverslips using Fluoromount G (Southern Biotech). Images were acquired at 60x on a Deltavision deconvolution microscope. Z-stacks of 5-10 0.2 micron steps were acquired and subjected to deconvolution analysis. 2-3 representative sections were selected and projections to 2D images were made and exported as tiff files.

### Immunofluorescence for adherent endothelial cells

Endothelial cells were plated on collagen 22mm round #1 German glass coverslips (Corning) in 6 well plates and were then fixed by adding 2% PFA for 15 min, rocking at room temperature and then rinsed. The adherent cells on the coverslips were then permeabilized and blocked in PBS with 0.2% Saponin (Sigma-Aldrich) and 5% normal goat serum (Invitrogen) for 15 min at room temperature, rocking. The coverslips were then transferred to a humidity chamber and incubated for 1 hour in PBS with 0.2% Saponin and 1% FBS with appropriate primary antibody. The coverslips were then washed, incubated with PBS and 0.2% Saponin + 1% FBS and secondary antibody for 30 min, washed and then mounted using Prolong Gold Antifade Reagent with DAPI (Cell Signaling Technology) overnight in the dark at room temperature. Six representative images for each experimental condition were analyzed for immunofluorescence. Images were obtained using the Bio Tek Lionheart FX Automated Microscope at 20x and were exported as tiffs. The following antibodies were used: mouse monoclonal antibody WT1 clone F-6, (1:200, Santa Cruz Biotechnology), and goat anti-mouse IgG conjugated to Alexa Flour 488 (Jackson ImmunoResearch) was used as a secondary antibody for immunofluorescence.

### Flow cytometry analysis

For cell surface staining, cells were incubated with the appropriate monoclonal antibodies for 30 min on ice in the dark, washed and incubated with secondary antibody reagents. Flow cytometry data were collected on a FACSCalibur (Beckton-Dickinson) and analyzed with FlowJo V 10.7 and 10.7.1 software. The following antibodies were used for flow cytometry: a monoclonal antibody against human HLA-A2 (clone BB7.2, ThemoFisher Scientific), conjugated with APC-eFlour 780, the human monoclonal antibody ESK1 against WT1 peptide/ HLA-A0201 complex ^22^, and an IgG2b isotype control (BD Biosciences).

### Statistical Analysis

All statistical analysis was performed using GraphPad Prism version 8.0 for Windows (GraphPad Software), where p-values <0.05 are considered significant. Experiments were done three times except where noted. Statistical significance was determined by Student’s t-test for unpaired individual comparisons. When 3 or more groups were analyzed, an ANOVA test was used. Linear and nonlinear regression analyses were used to fit curves.

## Results

### WT1 is overexpressed in KS and correlates with LANA expression

KS tumor biopsies from 303 HIV+ participants in the multicenter clinical trial^49^ were sectioned for H&E, and IHC for WT1 and LANA (**Figure 1A**). Double IHC for both revealed WT1 expression in LANA+ cells (**Figure 1A**). WT1 was expressed in areas involved by KS, but not in adjacent normal skin **(Figure 1B)**, which was quantified in 5 cases (p= 0.0004; **Supplemental Figure S1A**). Histopathologic classification into patch, plaque and nodular stages was perfomed^3^. Each case was analyzed by HALO software for WT1 expression to objectively derive the percentage of positive cells in the areas histologically involved by KS. We grouped cases based on the percentage of WT1 positive as: 1-20% (1+), 21-50% (2+), >50% (3+). Moderate to strong WT1 expression (>20% WT1 positive cells) was found in 64.7% of the 303 biopsies, and in 92.2% of nodular lesions. WT1 expression correlated with increasing histopathologic stage (p<0.0001; **Figure 1C-D**). Furthermore, WT1 expression was positively correlated with expression of LANA (p<0.0001; **Figure 1E**). This indicates that infected spindle cells express higher levels of WT1, and that the proportion of both WT1 and LANA positivity increase with histopathological stage. Additional validation cohorts were examined by IHC. Among 40 cases obtained in the US, a paired cohort with a range of histologies that were similar in number (patch, plaques, and nodules) from 15 HIV-positive and 15 HIV-negative patients showed significantly higher WT1 expression in the HIV-positive cohort (p=0.027) (**Figure 1F**). Another US cohort of 10 HIV-PLWH showed 2+ or 3+ WT1 expression in 8 of 10 cases (**Supplemental Figure S1B and S1C**). An additional cohort of PLWH from Uganda showed 2+ to 3+ WT1 expression in 18 of 20 cases (**Supplemental Figure S1B and S1C**).

**Figure 1.**
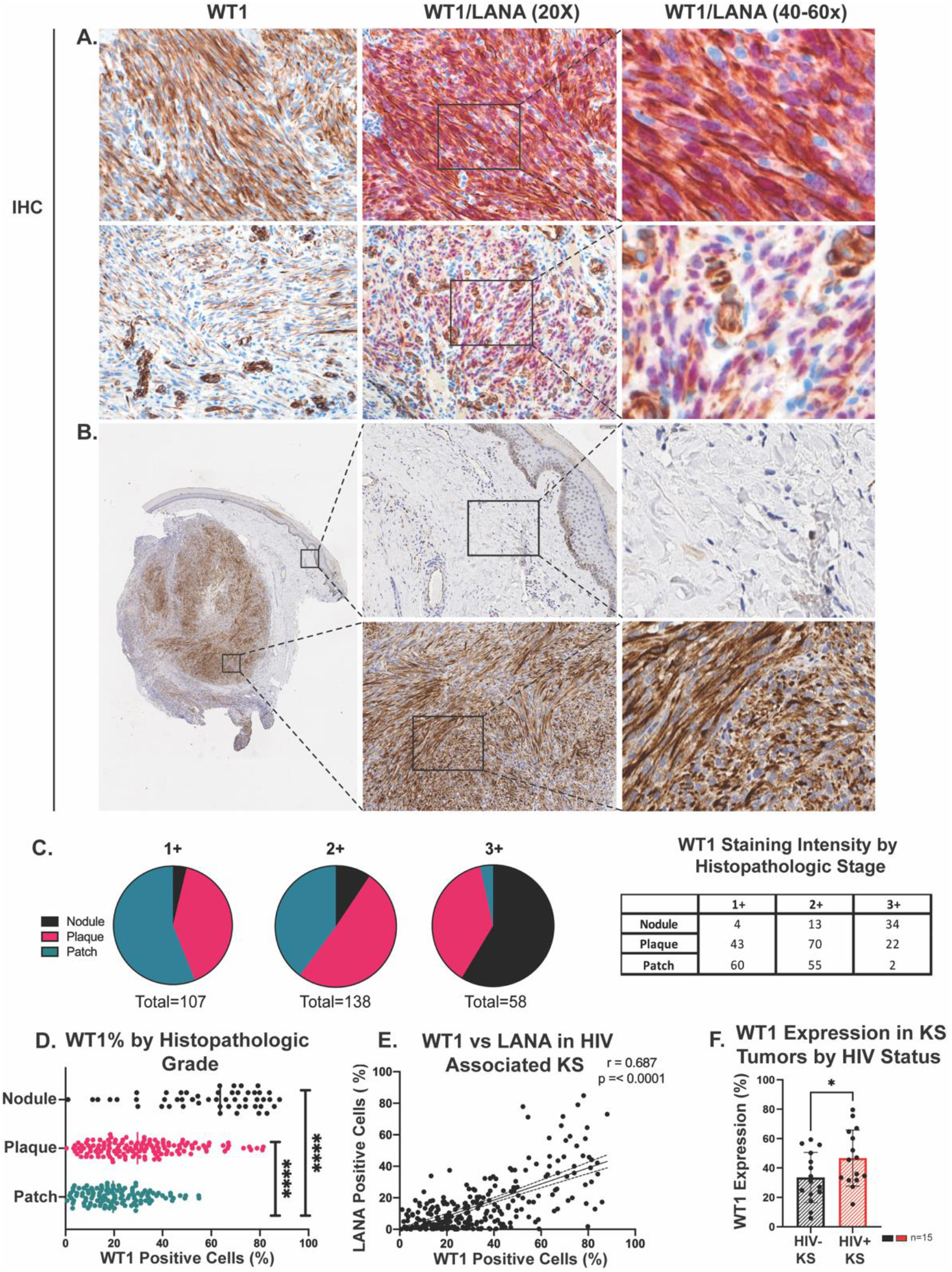
WT1 Expression in Kaposi Sarcoma. **A**. Two representative cases showing WT1 overexpression by IHC in KS (left panels), and by double IHC for WT1 (brown, nuclear and cytoplasmic) and KSHV LANA (red, nuclear) (middle panels, with zoom-in on right panels). Original magnification, 20X. **B**. Example of a KS nodule with adjacent skin tissue, showing a low power (4x, left panel) and higher power images (20x, middle panels) of uninvolved skin (top) and KS involved skin (bottom). Right panels show a zoom-in of the areas indicated. **C**. Pie charts of the IHC for WT1 in 303 cases from ACTG and AMC AIDS-KS trial AMC066/A5263 (NCT01435018), classified as 1+=1-20%, 2+= >20-50%, 3+=>50-100%, as determined by % positive cells by immunohistochemistry using quantitative image analysis. 2-3+ WT1 staining occurred in 64.7% of all cases and 92.2% of nodular lesions. **D**. Higher proportion of WT1-positive cells occurred with increased histopathologic grade demonstrated by scatter plot, patch vs. plaque p<0.0001, patch vs. nodule, p<0.0001, and plaque vs. nodule, p<0.0001 (using Tukey’s multiple comparison test). **E**. Proportion of WT1 expressing cells correlated significantly with proportion of LANA+ cells (r=0.687, p<0.0001), as determined by IHC and image analysis. **F**. WT1 expression examined in 15 tumor biopsies from HIV positive and 15 from HIV negative patients, demonstrating increased WT1 expression in the HIV positive group (p=.027).

### WT1 is upregulated by KSHV

After *in vitro* KSHV infection, WT1 mRNA and protein levels were increased in HUVEC, iLEC, HuARLT-1, and 293T-HEK. Peak upregulation occurred between 3 days and 6 days after establishment of KSHV **(Figure 2A and 2B)**. Infection of these cells was documented by IHC (Figure 2C) or GFP expression by KSHV-BAC-16 (**Figure 2D**). *In vitro* infection was variable, and usually occurred in less than 40% of the total cellular population, so levels of WT1 seen by western blot likely underestimate upregulation in the infected cells. In a telomerized endothelial cell line with a paired stably KSHV-infected counterpart (TIVE and TIVE-LTC), there were also higher WT1 levels in the infected cells. The WT1 band found to increase as a result of KSHV infection in western blots migrated near 62-68 kD, consistent with the cug-WT1 isoform previously identified as a major oncogenic isoform^16^.

**Figure 2.**
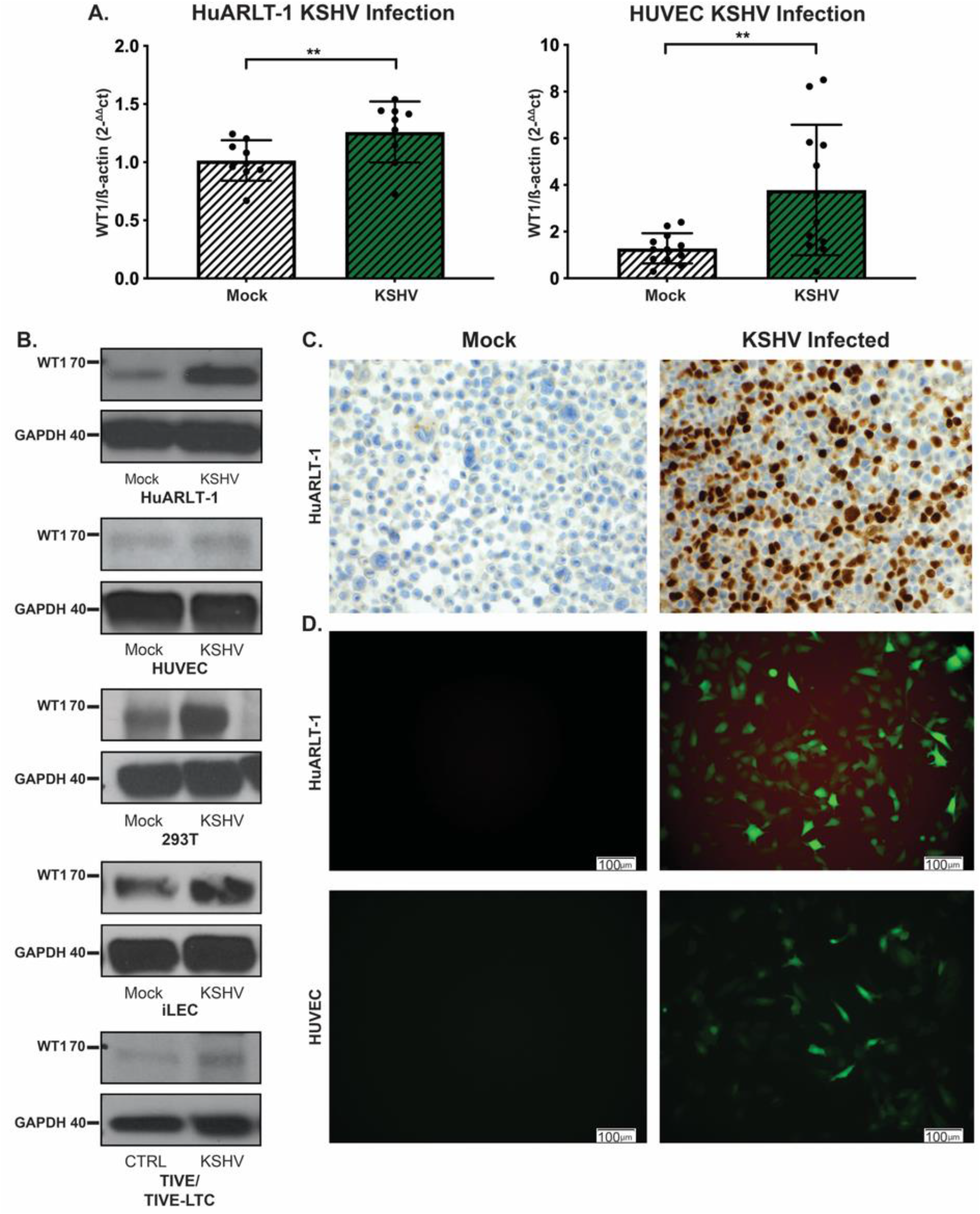
WT1 is upregulated in *in-vitro* de novo KSHV infection. **A**. qRT-PCR showing WT1 mRNA levels in mock vs. KSHV-BAC16 infection of HuARLT-1 and HUVEC endothelial cells, shown normalized to β-actin, are increased after KSHV infection. **B**. Western blots of WT1 protein (62-68kD size) in HuARLT-1, HUVEC, 293T, iLEC, and TIVE vs. TIVE-LTC showing higher levels in the setting of KSHV infection. Representative images are shown of at least three independent experiments. **C**. Mock vs. KSHV infection of HuARLT-1 cell blocks with IHC for LANA, documenting infection. **D**. Representative images obtained under fluoroscopy for GFP, indicative of KSHV infection in HuARLT-1 and HUVEC cells.

### The latent viral onco-protein vFLIP upregulates WT1 *in-vitro*

Amongst the KSHV proteins, vFLIP may have an impact on WT1 expression due to its ability to induce NFκB, a transcription factor known to regulate WT1 levels^33^. However, while vFLIP is a latent mRNA and vFLIP protein has been shown to be expressed in PEL cell lines during latency, the expression of vFLIP protein in KS lesions has not been previously documented, given the lack of robust anti-vFLIP antibodies useful for IHC. By applying an immunofluorescence method with tyramide amplification developed for this purpose, we found vFLIP expression in the cytoplasm of spindle cells of all four cases examined (**Figure 3A**), thus confirming that vFLIP is a latent viral protein expressed in KS.

**Figure 3.**
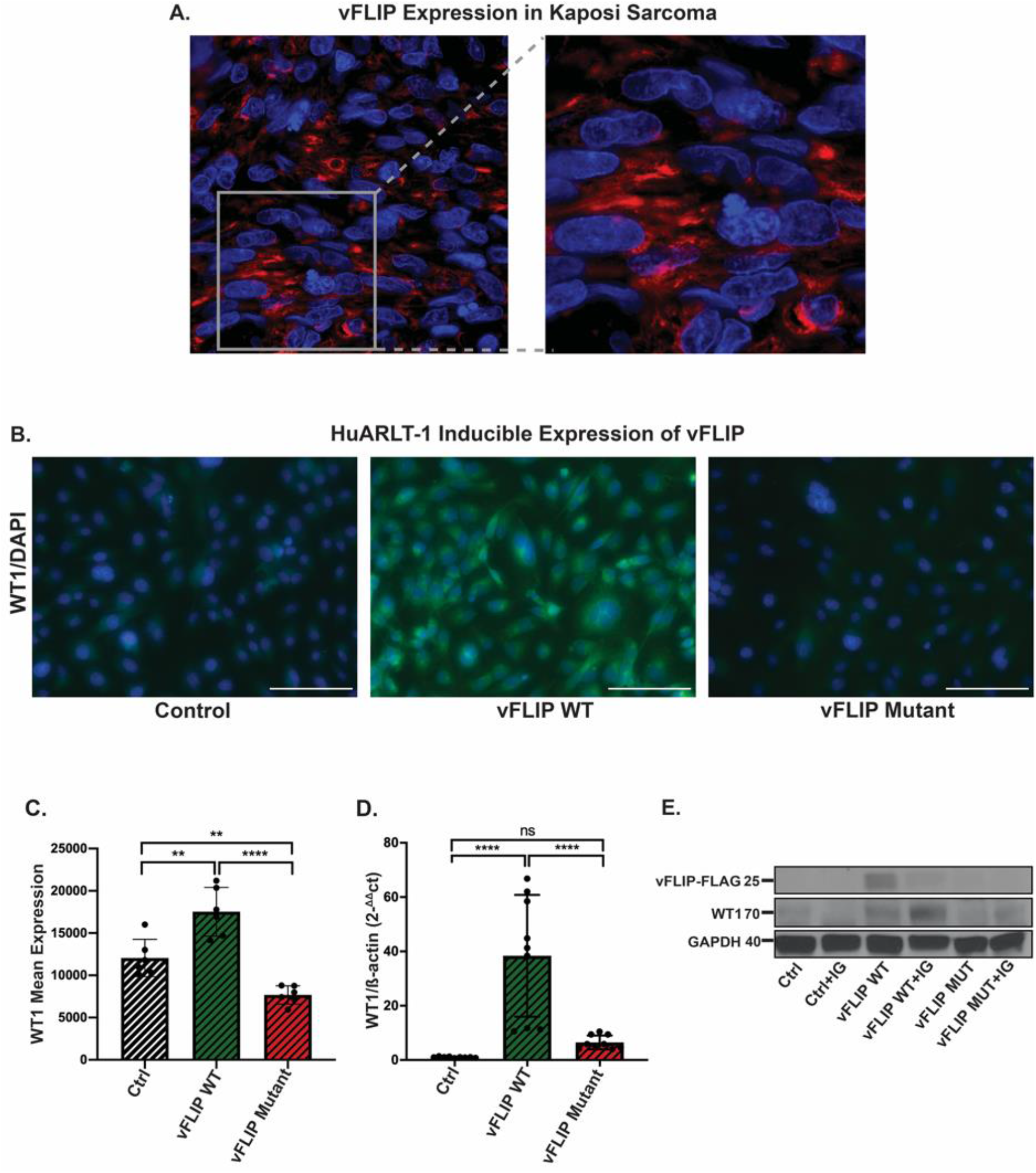
vFLIP upregulates WT1 *in vitro*. **A**. Immunofluorescence revealing vFLIP protein expression (red) in a representative primary KS biopsy showing cytoplasmic reactivity. **B**. vFLIP expression upregulates WT1, demonstrated by immunofluorescence (green) in HuARLT-1. A mutant vFLIP that is unable to bind IKK*γ* and induce NF𝒦B is defective in its ability to upregulate WT1. **C**. Quantitation of WT1 immunofluorescence upon vFLIP induction, performed by averaging the GFP fluorescence for six areas, showed increase with wild type vFLIP, but not with mutant vFLIP: control vs. vFLIP (p=0.0016), vFLIP vs. mutant vFLIP (p<0.0001). **D**. qRT-PCR for WT1 showed induction of WT1 mRNA with wild type vFLIP expression, which was minimal upon mutant vFLIP expression: control vs. vFLIP (p<0.0001) and vFLIP vs. mutant vFLIP (p<0.0001). **E**. Western blots show that WT1 protein is increased upon vFLIP-FLAG induction of expression in HuARLT-1 cells (cugWT1 ∼62-68kD). This increase was not seen when the mutant vFLIP-FLAG is expressed, which is also less stable than the wild type form, as seen with antibodies to FLAG. WT1 protein is further augmented in the presence of vFLIP induction and IFN-*γ* (IG).

To test the ability of vFLIP to affect WT1 expression, HuARLT-1 cell lines were transduced with a doxycycline inducible pLVX vFLIP-FLAG lentivirus. Stable lines were established with puromycin selection. WT1 was upregulated upon vFLIP induction, as determined by immunofluorescence (p=0.0016; **Figure 3B and 3C**). WT1 expression was also evaluated using a vFLIP NFκB dead mutant (vFLIP^AAA(58-60)^) that renders vFLIP unable to bind IKK*γ*. Wild type vFLIP showed higher WT1 induction than mutant vFLIP (p<0.0001; **Figure 3C**). This finding was confirmed at the RNA level by RT-PCR, and at the protein level by immunoblotting in HuARLT-1 cells, where WT1 levels were increased with vFLIP expression in both assays, but not with mutant vFLIP (**Figure 3D and 3E**). As expected, mutant vFLIP protein levels were lower, because the protein expression is dependent on binding to IKK*γ* which is disrupted. Similar to KSHV infection, upregulation of the cugWT1 ∼ 62-68kD band was seen in the presence of vFLIP. WT1 protein is further augmented in the presence of IFN-*γ* and wild type vFLIP induction, but not in mutant vFLIP-expressing cells. IFN-*γ* increases presentation of WT1 through upregulation of MHC class 1 molecules. These findings were validated in HUVEC cells, where vFLIP expression also resulted in increased WT1 levels (**Supplemental Fig 2A**). These findings indicate that KSHV-mediated WT1 upregulation is at least partly mediated by vFLIP (**Figure 3B-3E and Supplemental 2A and 2B**).

### WT1 is inversely correlated with T cells in the tumor microenvironment

Given that previous models suggest that WT1 may play a role in regulating the presence of immune cells in the tumor microenvironment^20^, the spatial relationship between expression of WT1 and LANA with immune infiltrates, including CD8+T, CD4+T cells, CD20+ B cells and plasma cells, was examined. In general, few B cells were seen, and plasma cells ranged from very abundant, sometimes in large clusters, to sparse. In general, CD8+ T cells were more abundant than CD4+ T cells. These were present in areas rich in inflammatory cells, which were usually concentrated at the margin of regions involved by KS (LANA+ and WT1+). This was more evident in nodular lesions, where KS areas are well defined (**Figure 4A**). CD8+ and CD4+ T cells were also present within areas containing sheets of spindle cells within KS nodules, but these were sparse and dispersed, rather than forming inflammatory aggregates seen at the periphery of these lesions. To formally assess a possible relationship between WT1 and CD8 expression in KS lesions we conducted image analysis using HALO software to create pseudocolor images of IHC for LANA, WT1, CD4 and CD8 (**Figure 4A**). Overall correlation analysis revealed an inverse relationship between WT1 expression and CD8+T cells (r=-0.2536; p<0.0001; **Figure 4B**). Thirteen nodules, eleven plaques and twelve patches were evaluable as sequential sections and analyzed by selecting five corresponding high LANA and high CD8+ T cell abundant areas for each lesion (**Figure 4C**). Focusing on areas with high WT1+ and LANA+ cells within KS nodules, there were notably lower percentages of CD8+T cells and CD4+T cells in these regions (LANA=60% vs. CD8=16%, p<0.001; LANA=60% vs. CD4=17%, p=0.0002; WT1=91% vs. CD8=16%, p<0.0001; and WT1=91% vs. CD4=17, p<0.0001), whereas in areas of low WT1+/LANA+ cells immediately adjacent to nodules, plaques and patches there was a higher percentage of CD8+T cells (CD8=67% vs LANA=4%; p<0.0001 and CD8=67% vs WT1=13.11%, p<0.0001) (**Figure 4D and Supplemental Table 2**). Taken together, these findings indicate that T cell-rich areas are outside the KS lesions, and are consistent with a relatively immunosuppressive immune environment in the immediate proximity of the KSHV-infected spindle cells.

**Figure 4.**
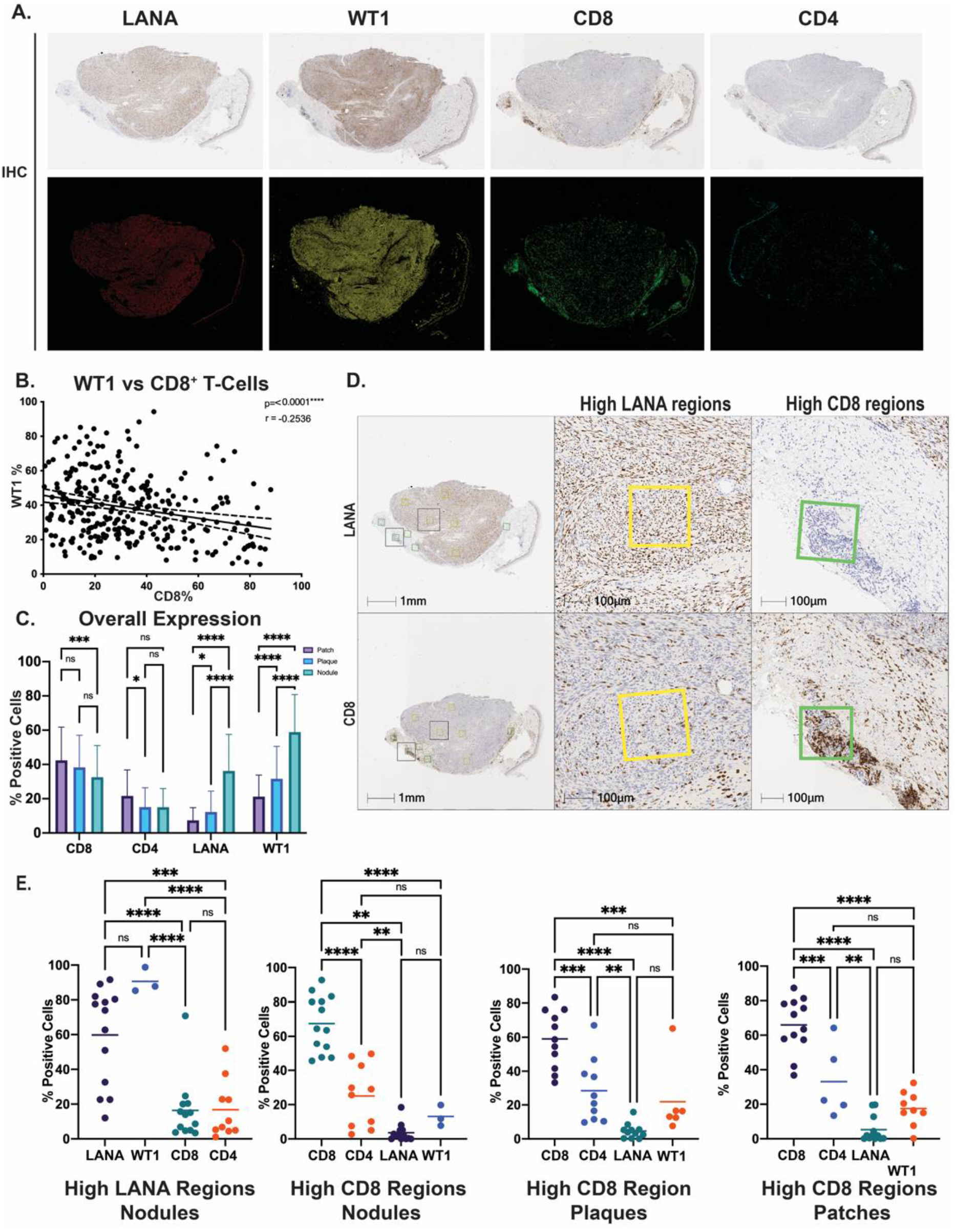
WT1 and LANA expression correlate with decreased T cell infiltrates. **A**. Using HALO analysis software of KS tumors from AMC066/A5263 clinical trial, pseudocolor images were created for IHC of LANA, WT1, CD8, and CD4 cells. **B**. WT1 expression in all cases was inversely correlated with the number of CD8+T cells. **C**. Assessment of cellular populations according to histological stage showed lower CD8 and CD4 T cells with higher histological grade, and the reverse for WT1 and LANA. **D**. Clusters of CD8+ cells were seen at the periphery of nodular KS lesions. Areas with numerous LANA+ cells were selected and quantified for CD8 positivity in a sequential tissue section, and vice versa. **E**. This spatial analysis was performed for WT1, LANA, CD4+T cells and CD8+T cells in areas of high LANA or high CD8 in nodular lesions, and high CD8 +T cell regions in corresponding regions for WT1/LANA/CD4 in nodules, plaques and patches.

### WT1 is a potential target for immunotherapy in Kaposi sarcoma

The effect of KSHV infection on binding of ESK1 antibodies, which recognize WT1 peptides presented on HLA-A0201, was tested by transducing HuARLT-1 with HLA-A0201, followed by KSHV infection. Subsequent incubation with ESK-1 showed 23% positivity in mock cells vs 28% in KSHV infected cells (p=0.0465; **Figure 5A)** of which approximately 30% of cells were KSHV infected by GFP expression. In addition, there was higher ESK1 binding upon vFLIP induction of HuARLT-1 cells, which increased from 7% to 19% (p= 0.0004) (**Figure 5B**), consistent with the hypothesis that vFLIP expression alone upregulates WT1 and increases ESK-1 binding.

**Figure 5.**
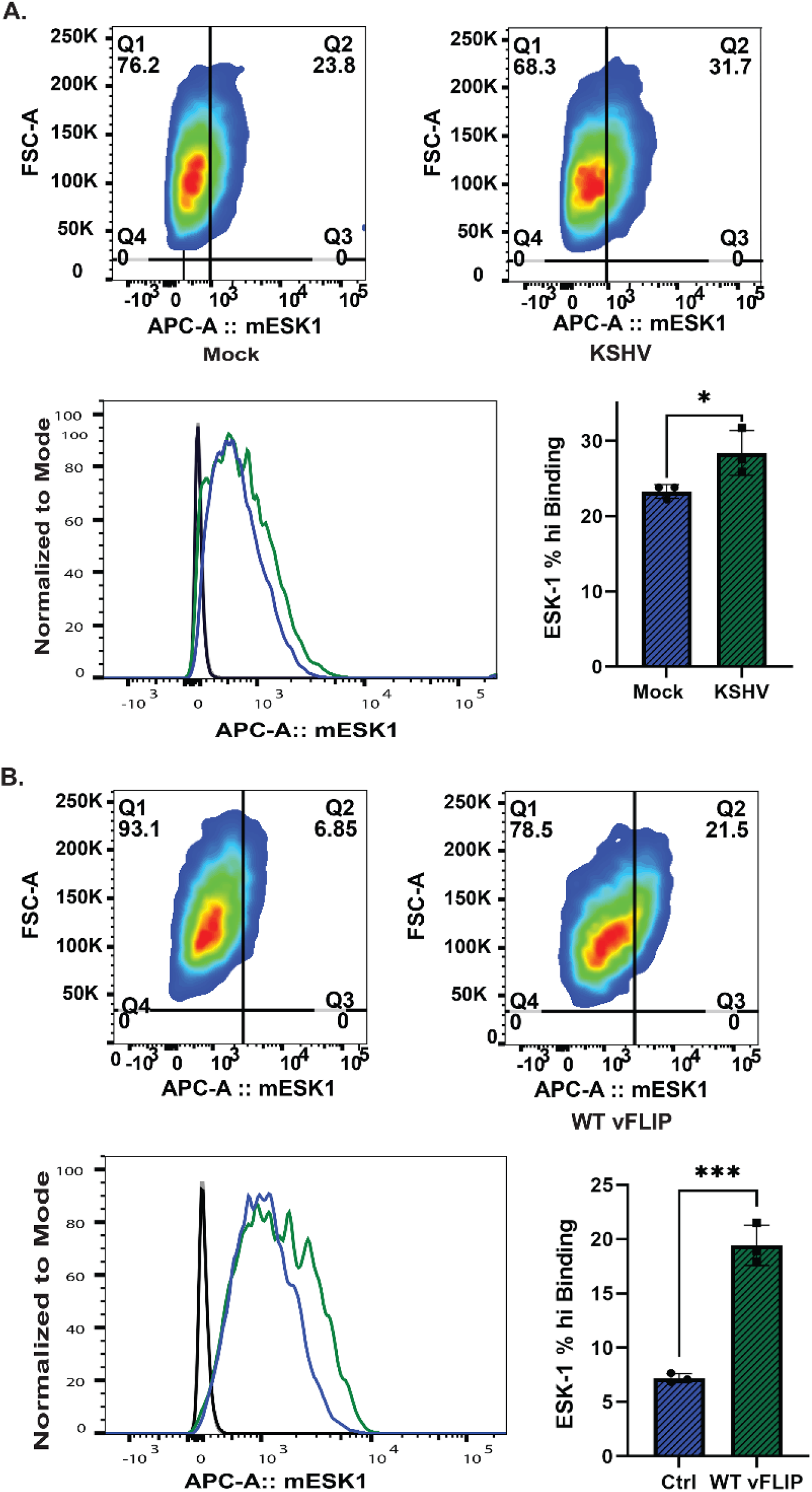
WT1 is a potential target for immunotherapy in Kaposi sarcoma. **A**. ESK1 binding assays with HLA-A0201 transduced HuARLT-1 cells at 6 days post KSHV infection (green) or mock infection (blue) and ESK-1 antibody incubation compared to IgG isotype controls (black line), assessed through flow cytometry. **B**. ESK-1 binding assays performed with HLA-A0201 transduced HuARLT-1 cells as control (blue) vs. vFLIP (green) at 96 hours of doxycycline induction and ESK-1 antibody incubation compared to IgG isotype controls (black line), assessed through flow cytometry.

## Discussion

This study shows that WT1 is highly expressed in the vast majority of Kaposi sarcoma lesions. Importantly, expression is higher in cases with histologically advanced KS. Our findings also show significantly higher WT1 expression in HIV-associated KS compared to HIV-negative KS lesions, even when paired with histologies similar in number. This observation raises the possibility that HIV may affect WT1 expression directly or indirectly. Clearly evident from our studies is that KSHV can upregulate WT1 in endothelial cells. KSHV infection *in vitro* increases WT1, and in particular the 62-68kD isoform consistent with an oncogenic form of this protein, also called ‘cugWT1’. In KS lesions, KSHV is largely latent, expressing only a handful of viral gene products. Here we confirm that one of these, vFLIP, is in fact expressed in KS lesions and demonstrate that this viral gene alone also increases WT1 expression, indicating that this viral protein is at least partly responsible for KSHV-mediated WT1 upregulation.

Analysis of KS tumor specimens also revealed that there is an important microenvironmental impact on WT1 expression. While upregulation of WT1 in KSHV-infected cells in vitro was consistent among multiple experiments and different endothelial cell types, WT1 levels appear to be much higher *in vivo*. Moreover, WT1 expression in tissue biopsies significantly correlated with the expression of viral latent oncoprotein LANA, and inversely correlated with T cells within the tumor and surrounding stromal tissue. Furthermore, the low T cell infiltration noted in areas of high WT1 and high LANA regions suggest that WT1 may contribute to the immunosuppressive functions of KSHV in creating an immunosuppressive tumor microenvironment. In fact, one study found that knockout of WT1 in mouse endothelial, hematopoietic and myeloid suppressor cells (MDSCs), led to decreased tumor growth and metastases, and that WT1 was critical to recruiting MDSCs to suppress T cell immune responses^20^. In addition, our observations are consistent with a recent study that demonstrated decreased immune infiltrates in areas of KS lesions showing abundant KSHV-infected cells compared to surrounding areas^43^. Nevertheless, while both CD4+ and CD8+ T cells are present among the spindle cells, their functional phenotypes remained to be characterized.

Thus, ongoing studies are aimed at characterizing the immune components of KS lesions in depth, including further characterization of T cells subsets (i.e. activation, exhaustion, Tregs, memory), as well as other cells, such as tumor associated macrophages (TAMs) and MDSCs. Our lab has previously generated transgenic mice expressing vFLIP in endothelial cells, which revealed remodeling of myeloid differentiation with M1 toward M2 polarization, and expansion of MDSCs and TAMs. These *in vivo* observations are consistent with a model whereby vFLIP induces WT1, which in turn recruits MDSCs to the tumor microenvironment. Furthermore, WT1 has been shown to induce a number of other factors that can promote tumorigenesis and angiogenesis and have been shown to be expressed by KS spindle cells or KSHV infected endothelial cells in culture, including MMP9, VEGF and PDGF receptors^44-46^.

We also investigated whether a TCR mimic antibody against WT1 and HLA-A0201 would bind endothelial cells with increased WT1 expression as a result of KSHV infection or vFLIP induction. We found that both conditions increased ESK-1 binding. These findings suggest that WT1 directed immunotherapy, either with this antibody, or with bi-specific T cell-engaging antibodies (BiTEs)^47^ or anti-WT1 T cells or^27^ CAR T cells,^48, 49^ may have therapeutic potential in KS. Other immunotherapeutic approaches include WT1-directed peptide vaccines, which have been studied in phase 1 and phase 2 trials for hematologic and solid malignancies^25, 50^. Given that WT1 peptide vaccines have been found to be safe and induced clinical and immunological responses^23, 25^, this approach may be attractive for use as a therapeutic vaccine against KS, especially in chemotherapy-resistant or recurrent cases.

We propose a model of WT1 mediated tumorigenesis in Kaposi sarcoma in which following KSHV infection, latent viral genes are expressed, including vFLIP, which activate NFκB, leading to upregulation of the oncogenic cugWT1 isoform. In turn, WT1 expression, in conjunction with expression of other viral genes, leads to an immunosuppressive and angiogenic tumor microenvironment. We propose that given the high WT1 expression in the majority of KS lesions, immunotherapy directed against WT1 might reverse the immunosuppressive KS microenvironment. Lastly, WT1 expression may serve as a biomarker to identify cases most likely to respond to WT1 immunotherapy, which may be especially important in cases of advanced KS that disproportionately affect PLWH.

## Data Availability

Data available within the article or its supplementary materials.

## Acknowledgements

This investigation was supported by grant KL2TR002385 of the Clinical and Translation Science Center at Weill Cornell Medical College to AM and NIH R01 R01CA CA250074 to EC. Clinical trial AMC066/A5263 was supported by AMC grant U01 CA121947 from the National Cancer Institute and the ACTG awards UM1 AI068634, UM1 AI068636, and UM1 AI106701 from the National Institute of Allergy and Infectious Diseases of the National Institutes of Health. The content is solely the responsibility of the authors and does not necessarily represent the official views of the National Institutes of Health. We thank Dr. Jae Jung from the Keck School of Medicine, University of Southern California for generously providing the recombinant KSHV BAC16 constructs. We thank Dr. Michael Sturzl from University of Erlangen-Nuremberg, Germany for providing the KSHV vFLIP rat monoclonal antibody. We thank Dr. Annamalai Selvakumar, PhD, and Dr. Richard O’Reilly for providing the HLA-A0201 plasmid vector. We thank Jouliana Sadek and Jessica Osborn, and other current and previous members of the Weill Cornell Medicine-Cesarman laboratory for critical discussion and feedback. Project support for this research was provided in part by the Center for Translational Pathology at the Department of Pathology and Laboratory Medicine, Weill Cornell Medicine.

---

**Supplemental Table 1.**
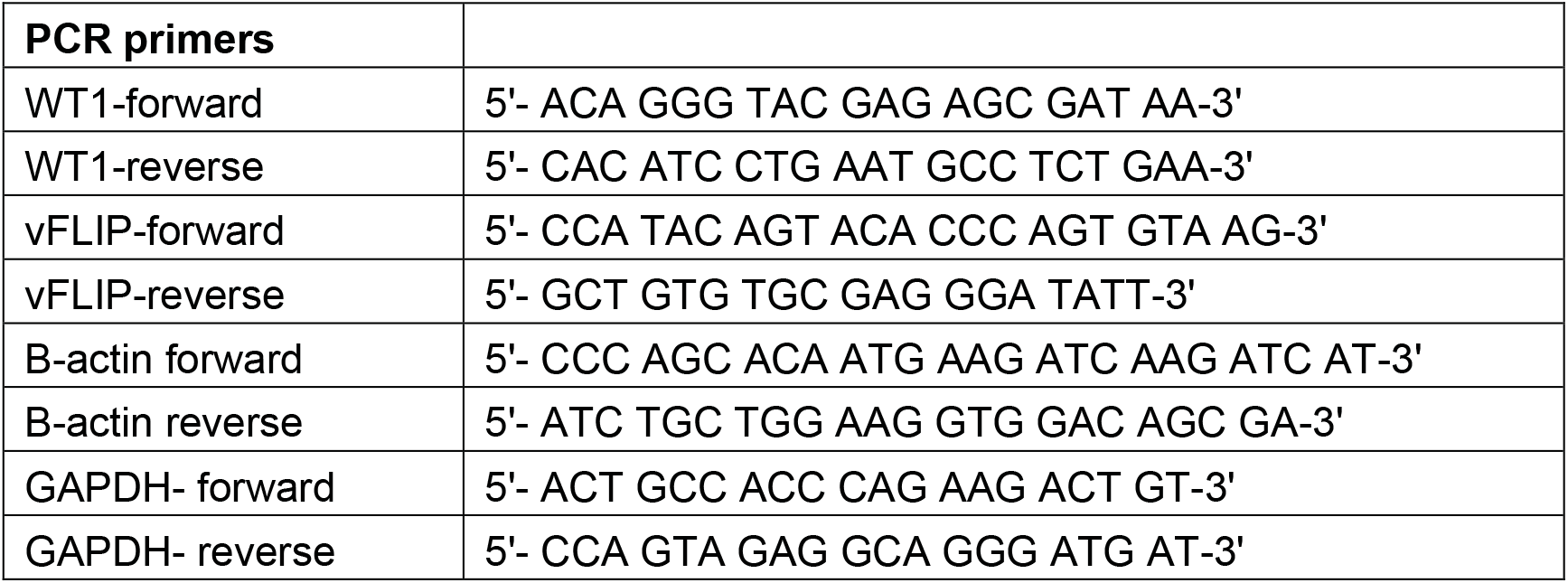
RT-PCR Primers.

**Supplemental Table 2.** Correlation analysis of Immune Infiltrates and WT1/LANA in KS Nodules, Plaques and Patches.

| **Ordinary one-way ANOVA of High LANA Nodules** |  |  |  |  |  |  |
| --- | --- | --- | --- | --- | --- | --- |
| Tukey's multiple comparisons test | Mean Diff. | 95.00% CI of diff. | Below threshold | Summary | Adjusted P Value |  |
| CD8 vs. CD4 | 42.34 | 26.79 to 57.90 | Yes | **** | <0.0001 | A-B |
| CD8 vs. LANA | 63.79 | 49.29 to 78.30 | Yes | **** | <0.0001 | A-C |
| CD8 vs. WT1 | 54.31 | 30.62 to 78.00 | Yes | **** | <0.0001 | A-D |
| CD4 vs. LANA | 21.45 | 5.892 to 37.01 | Yes | ** | 0.0037 | B-C |
| CD4 vs. WT1 | 11.96 | -12.38 to 36.31 | No | ns | 0.5535 | B-D |
| LANA vs. WT1 | -9.485 | -33.17 to 14.21 | No | ns | 0.7040 | C-D |
| Test details | Mean 1 | Mean 2 | Mean Diff. | SE of diff. | n1 | n2 |
| CD8 vs. CD4 | 67.42 | 25.07 | 42.34 | 5.768 | 13 | 10 |
| CD8 vs. LANA | 67.42 | 3.626 | 63.79 | 5.379 | 13 | 13 |
| CD8 vs. WT1 | 67.42 | 13.11 | 54.31 | 8.784 | 13 | 3 |
| CD4 vs. LANA | 25.07 | 3.626 | 21.45 | 5.768 | 10 | 13 |
| CD4 vs. WT1 | 25.07 | 13.11 | 11.96 | 9.028 | 10 | 3 |
| LANA vs. WT1 | 3.626 | 13.11 | -9.485 | 8.784 | 13 | 3 |
| **Ordinary one-way ANOVA of Nodule High CD8 Regions** |  |  |  |  |  |  |
| Tukey's multiple comparisons test | Mean Diff. | 95.00% CI of diff. | Below threshold | Summary | Adjusted P Value |  |
| CD8 vs. CD4 | 30.66 | 11.90 to 49.42 | Yes | *** | 0.0005 | A-B |
| CD8 vs. LANA | 54.42 | 36.11 to 72.73 | Yes | **** | <0.0001 | A-C |
| CD8 vs. WT1 | 37.09 | 15.30 to 58.88 | Yes | *** | 0.0003 | A-D |
| CD4 vs. LANA | 23.76 | 5.000 to 42.52 | Yes | ** | 0.0085 | B-C |
| CD4 vs. WT1 | 6.431 | -15.74 to 28.60 | No | ns | 0.8614 | B-D |
| LANA vs. WT1 | -17.33 | -39.12 to 4.462 | No | ns | 0.1588 | C-D |
| Test details | Mean 1 | Mean 2 | Mean Diff. | SE of diff. | n1 | n2 |
| CD8 vs. CD4 | 59.01 | 28.35 | 30.66 | 6.946 | 11 | 10 |
| CD8 vs. LANA | 59.01 | 4.588 | 54.42 | 6.779 | 11 | 11 |
| CD8 vs. WT1 | 59.01 | 21.92 | 37.09 | 8.068 | 11 | 6 |
| CD4 vs. LANA | 28.35 | 4.588 | 23.76 | 6.946 | 10 | 11 |
| CD4 vs. WT1 | 28.35 | 21.92 | 6.431 | 8.209 | 10 | 6 |
| LANA vs. WT1 | 4.588 | 21.92 | -17.33 | 8.068 | 11 | 6 |
| **Ordinary one-way ANOVA of Plaque High CD8 Regions** |  |  |  |  |  |  |
| Tukey's multiple comparisons test | Mean Diff. | 95.00% CI of diff. | Below threshold | Summary | Adjusted P Value |  |
| CD8 vs. CD4 | 30.66 | 11.90 to 49.42 | Yes | *** | 0.0005 | A-B |
| CD8 vs. LANA | 54.42 | 36.11 to 72.73 | Yes | **** | <0.0001 | A-C |
| CD8 vs. WT1 | 37.09 | 15.30 to 58.88 | Yes | *** | 0.0003 | A-D |
| CD4 vs. LANA | 23.76 | 5.000 to 42.52 | Yes | ** | 0.0085 | B-C |
| CD4 vs. WT1 | 6.431 | -15.74 to 28.60 | No | ns | 0.8614 | B-D |
| LANA vs. WT1 | -17.33 | -39.12 to 4.462 | No | ns | 0.1588 | C-D |
| Test details | Mean 1 | Mean 2 | Mean Diff. | SE of diff. | n1 | n2 |
| CD8 vs. CD4 | 59.01 | 28.35 | 30.66 | 6.946 | 11 | 10 |
| CD8 vs. LANA | 59.01 | 4.588 | 54.42 | 6.779 | 11 | 11 |
| CD8 vs. WT1 | 59.01 | 21.92 | 37.09 | 8.068 | 11 | 6 |
| CD4 vs. LANA | 28.35 | 4.588 | 23.76 | 6.946 | 10 | 11 |
| CD4 vs. WT1 | 28.35 | 21.92 | 6.431 | 8.209 | 10 | 6 |
| LANA vs. WT1 | 4.588 | 21.92 | -17.33 | 8.068 | 11 | 6 |
| **Ordinary one-way ANOVA of Patch High CD8 Regions** |  |  |  |  |  |  |
| Tukey's multiple comparisons test | Mean Diff. | 95.00% CI of diff. | Below threshold | Summary | Adjusted P Value |  |
| CD8 vs. CD4 | 32.84 | 13.79 to 51.90 | Yes | *** | 0.0003 | A-B |
| CD8 vs. LANA | 60.67 | 46.05 to 75.28 | Yes | **** | <0.0001 | A-C |
| CD8 vs. WT1 | 48.35 | 32.57 to 64.14 | Yes | **** | <0.0001 | A-D |
| CD4 vs. LANA | 27.82 | 8.767 to 46.88 | Yes | ** | 0.0021 | B-C |
| CD4 vs. WT1 | 15.51 | -4.457 to 35.48 | No | ns | 0.1742 | B-D |
| LANA vs. WT1 | -12.31 | -28.10 to 3.474 | No | ns | 0.1715 | C-D |
| Test details | Mean 1 | Mean 2 | Mean Diff. | SE of diff. | n1 | n2 |
| CD8 vs. CD4 | 65.93 | 33.09 | 32.84 | 7.056 | 12 | 5 |
| CD8 vs. LANA | 65.93 | 5.263 | 60.67 | 5.412 | 12 | 12 |
| CD8 vs. WT1 | 65.93 | 17.58 | 48.35 | 5.845 | 12 | 9 |
| CD4 vs. LANA | 33.09 | 5.263 | 27.82 | 7.056 | 5 | 12 |
| CD4 vs. WT1 | 33.09 | 17.58 | 15.51 | 7.394 | 5 | 9 |
| LANA vs. WT1 | 5.263 | 17.58 | -12.31 | 5.845 | 12 | 9 |

**Note:** Multiple Comparisons analysis using One way ANOVA for high LANA regions in nodules, and high CD8+T cell regions in nodules, plaques and patches.

**Supplemental Figure S1.**
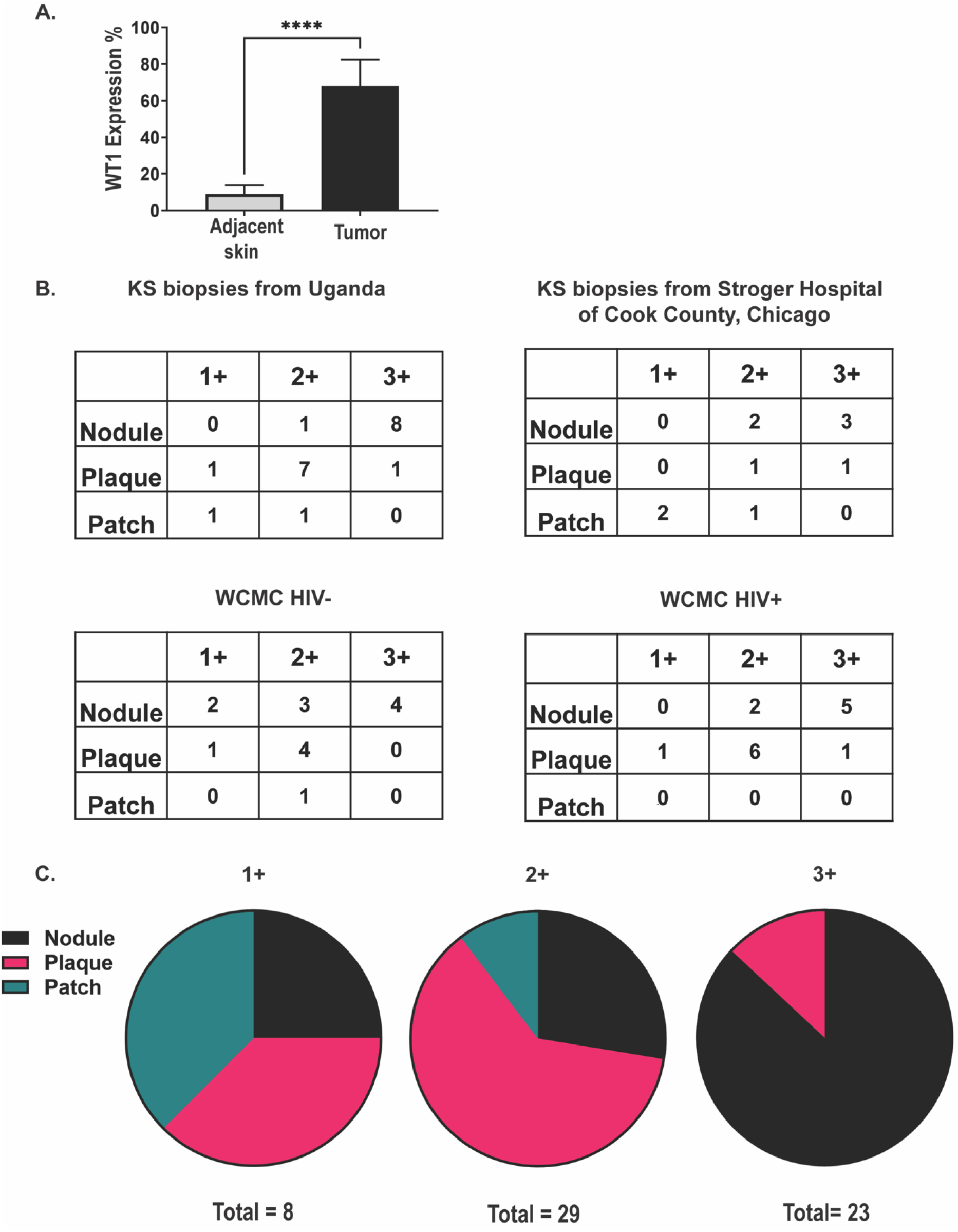
WT1 expression in vivo, Additional Cohorts. **A**. Five KS tissue biopsies were quantified for WT1 in the lesional tissue and adjacent normal subcutaneous tissue, demonstrating significant differential WT1 expression in KS compared to normal skin (p= 0.0004). **B**. Additional cohorts were examined for WT1 IHC, including in 10 cases from Stroger Hospital of Cook County, 20 cases from Uganda, and 30 cases from New York, that included KS from HIV+ and HIV– individuals. These cases were evaluated as weak (1+), Moderate (2+), Strong (3+) by microscopic examinations. **C**. The data from these additional cohorts is shown in pie charts, indicating that WT1 expression is higher with more advanced histopathological stages.

**Supplemental Figure S2.**
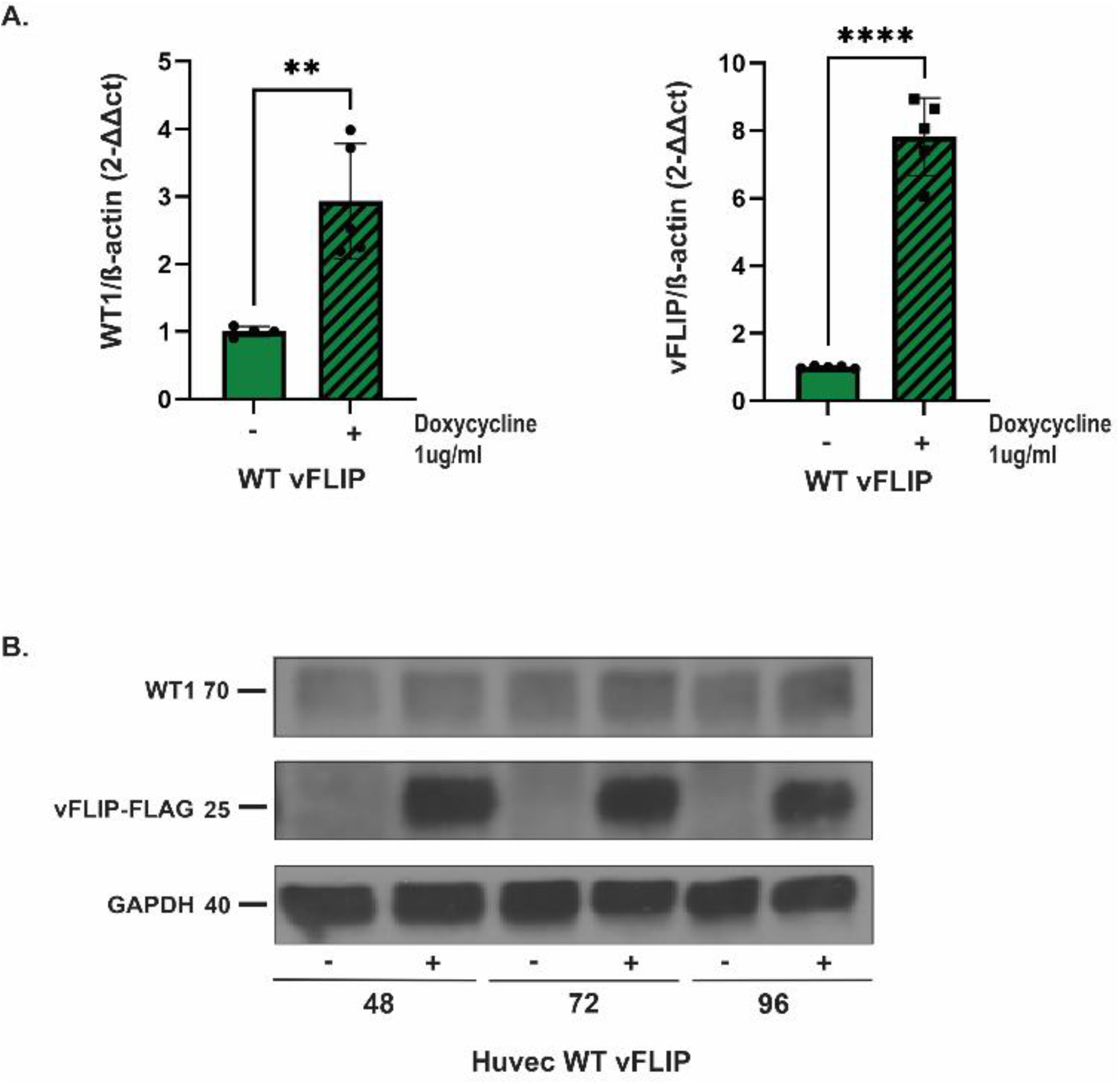
WT1 is upregulated in vitro upon induction of vFLIP expression in HUVECs. **A**. qRT-PCR showed WT1 upregulation (left) with vFLIP expression (right) in primary HUVEC cells at 96 hours of vFLIP induction. **B**. Western blots show a time course after induction of vFLIP-FLAG expression, indicating increasing WT1 levels corresponding with vFLIP expression.

**Supplemental Figure S3.**
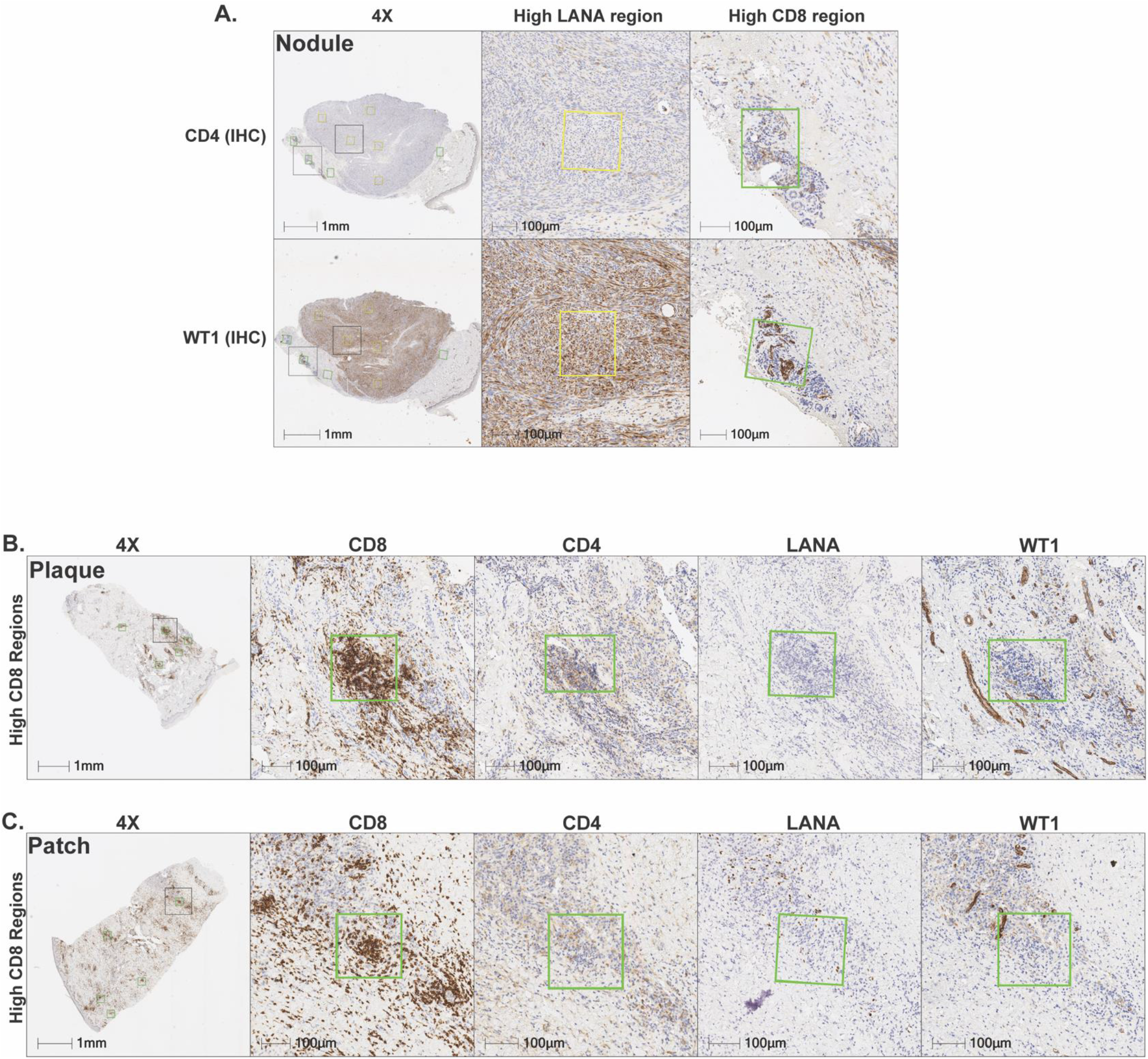
Inverse correlation of Immune Infiltrates and WT1/LANA in KS Nodules, Plaques and Patches. **A**. Representative detailed HALO analysis of both high LANA and high CD8+T cell regions, on sequential sections of the nodule from Fig. 4A, showing immunohistochemistry (IHC) for CD4+T cells and WT1. **B**. Representative detailed HALO analysis of high CD8+T cell regions with corresponding sequential sections for CD4+ T cells, LANA, and WT1 positive cells for plaque and patches.

